# Translation and psychometric validation of the Mental Illness: Clinicians’ Attitudes Scale (MICA-4) to assess attitudes of primary care physicians in Pakistan

**DOI:** 10.64898/2026.03.14.26347350

**Authors:** Noor ul Ain Muneeb, Asma Nisa, Asma Humayun

## Abstract

**Background:** Negative and stigmatizing attitudes towards people suffering from mental disorders among healthcare providers often act as a barrier to mental healthcare access. To assess these attitudes in primary care physicians (PCPs), a robust, culturally tailored psychometric tool is crucial. This study aimed to translate and psychometrically validate the MICA-4 to assess negative attitudes among PCPs in Pakistan.

**Methods:** We recruited two independent samples of PCPs (n=191, n=329) using non-probability sampling. Three bilingual mental health professionals forward-translated the scale, which was then independently reviewed and back-translated. Cognitive interviews were conducted (n=15 PCPs) to assess comprehension and clarity, for the final version to be used in the study. EFA was conducted on Sample 1 to examine the underlying factor structure of the Urdu MICA-4 items. CFA was then performed on Sample 2 to cross-validate the factor structure identified in Sample 1. Internal consistency and convergent validity were also assessed.

**Results:** A three-factor solution was retained, including Views (seven items), reflecting clinicians’ general evaluative perspectives toward mental illness and professional roles; Stereotypes (five items) representing generalized beliefs and disclosure-related concerns regarding individuals with mental illness, and Stigma (three items) capturing social distancing and perceived threat-related attitudes. The Comparative Fit (CFI = .958) and the Tucker-Lewis Index (TLI = .946) indicated good fit. Three items (9, 13, and 12) were removed due to weak loadings (< .40). Composite reliability (ω) indicated adequate internal consistency for the Views (ω = .70) and Stereotypes (ω = .74) factors, and lower for stigma (ω ≈ .53). Convergent validity was modest (.40 to .44).

**Conclusion:** The findings support the cautious use of Urdu MICA-4 in Pakistani primary care settings. The variability in the factor structure of the scale across cultures raises a practical implication for its dissemination. When item-level instability repeatedly emerges across contexts, permitting limited, evidence-based refinement may strengthen measurement stability and comparability, as well as its reliability in diverse healthcare settings.

## INTRODUCTION

With an estimated global burden of 12%, the prevalence of mental disorders has grown by 48.1 % in the last few decades (Arias et al., 2022; Global Burden of Disease, 2022). Despite this, the treatment gap shows that over 90% of people in low- and middle-income countries (LMICs) do not have access to evidence-based care (World Health Organization, 2022).

Some of the challenges to address this gap are the barriers rooted in structural inadequacies, as well as the pervasive influence of stigma (Corrigan et al., 2014; Vistorte et al., 2018). These barriers hamper access to services and lead to delayed help-seeking, suboptimal treatment, and risk of human rights violations (Javed et al., 2021). A comparative study revealed that only 12 to 15% of respondents from LMICs, compared to 45 to 51% in HICs, believe that mental illness is a health condition, much like a physical illness (Seeman et al., 2016). This disconnect in LMICs is reinforced by low mental health literacy and pervasive beliefs attributing mental illnesses to supernatural or religious causes, which often lead to faith healing being the primary treatment choice for mental disorders (Shah et al., 2019). The healthcare providers (HCPs) often believe that people with mental illness are dangerous, morally weak, or unpredictable (Laraib et al., 2018; Naeem et al., 2006). As a result of these negative attitudes, HCPs experience discomfort in interacting with patients and do not have conviction in therapeutic interventions which undermine treatment outcomes (Vistorte et al., 2018).

Primary care physicians (PCPs) are the first and often only point of contact for many individuals with mental illness in Pakistan (Hussain et al., 2018). There is neither adequate exposure to mental health training as part of their pre-service education, nor any regulatory framework exists mandating this training as part of their continual professional development (Farazdaq et al., 2022). Their clinical workload also discourages additional training or active engagement with people with mental healthcare needs. Unfortunately, these stigmatizing attitudes in clinicians reinforce the barriers that prevent patients from seeking professional help (Phelan et al., 2023; Vistorte et al., 2018). These also influence treatment adherence and mistrust in mental health services (Faregh et al., 2019).

Stigma is usually seen as abstract construct which is difficult to quantify or intervene upon without reliable and valid measures (Mitchell et al., 2018). Therefore, a robust psychometric and culturally tailored tool is important to assess negative attitudes towards mental illness in clinicians before these could be addressed. While community attitude measures like the Community Attitudes Toward the Mentally Ill (CAMI) and has been widely used, it is designed for community settings, is lengthy, and might not capture nuanced and complex clinical perspectives. It has also been suggested that the methodological quality of studies validating CAMI was low, and showed poor construct validity when applied to HCPs (Högberg et al., 2008; Kassam et al., 2010; Sanabria-Mazo et al., 2023). Similarly, Opinions About Mental Illness (OMI) is validated on university students and is also not generalizable to HCPs (Raj et al., 2023; Todor, 2013).

MICA-4 addresses these gaps and has been designed to assess stigma among HCPs to support training and policy efforts aimed at reducing stigma and barriers in healthcare access (Gabbidon et al., 2013; Clement et al., 2015).

It has also demonstrated sound psychometric properties and has been successfully adapted in both diverse high-and low-income countries, affirming its cultural adaptability and psychometric soundness across varied healthcare systems (Fernandes et al., 2024; Mısır et al., 2024; Vistorte et al., 2023). However, the dimensionality and factor structure of the scale vary, reflecting diversity in attitudes towards mental illness in different cultures (Fernandes et al., 2024; Gabbidon et al., 2013; Mısır et al., 2024; Vistorte et al., 2023).

A number of these instruments have been applied to assess the attitude of the community and HCPs in Pakistan (Haddad et al., 2016; Husain et al., 2020; Laraib et al., 2018; Talib et al., 2024), including MICA-4 (Khan et al. 2023). However, none of these instruments were contextualized and are likely to compromise internal validity and mislead results. Given the lack of a validated tool to measure stigmatizing attitudes among primary care physicians (PCPs), we aimed to translate and psychometrically validate the MICA-4 for Pakistan.

## 2. METHODS

### 2.1 Sample and Study Design

We employed a cross-sectional design and recruited two independent samples of primary care physicians (PCPs) using non-probability sampling techniques. We disseminated the survey through professional medical associations and postgraduate training networks. We also invited the participants to share the survey link with other eligible colleagues. In accordance with methodological guidelines, we recruited a total of 191 participants (Sample 1) for exploratory factor analysis (EFA) for an adequate ratio (five participants per item is generally considered adequate).

For Sample 2, we recruited an independent sample of 329 PCPs using purposive sampling for confirmatory factor analysis (CFA) to ensure stable parameter estimation and reliable model fit indices (sample exceeding 200 participants is recommended). Both samples met or exceeded these. The recruitment criteria are mentioned in *supplementary material A*.

We clearly explained the purpose and procedures of the study to all participants, and obtained written informed consent before survey completion. Both samples participated voluntarily.

### 2.2 Measures

Participants completed a brief demographic questionnaire assessing age, gender, level of education, years of clinical experience, and work sector. We selected the Mental Illness: Clinicians’ Attitudes Scale (MICA-4) which is a 16-item self-report scale designed to assess clinician’s attitudes towards individuals with mental health conditions. Items are rated on a 6-point Likert scale ranging from 1 = *strongly agree* to 6 = *strongly disagree*, with ten reverse-scored items.

### 2.3 Translation of MICA-4

We reviewed the instructions by the author for the MICA-4 scale translation ^1^ and found these aligned with the World Health Organization for the translation protocol (2020).

For forward translation, our team consisted of three bilingual mental health professionals (two clinical psychologists and one psychiatrist) with over ten years of clinical experience and prior experience in translating psychological instruments. Translators were instructed to prioritize conceptual equivalence and clarity for the intended audience. The forward translations were reviewed by an expert panel comprising a psychiatrist and a PhD researcher. The panel resolved discrepancies through consensus and finalized the Urdu version, ensuring conceptual accuracy and clarity while retaining the meaning of the original items.

This version was independently back-translated into English by a bilingual clinical psychologist. The back-translated version was compared with the original instrument to verify conceptual consistency.

Cognitive interviews were then conducted with 15 PCPs (9 males, 6 females) to assess comprehension and clarity. Participants were asked to explain their understanding of each item and identify any ambiguous terms. Minor linguistic refinements were made to improve clarity without altering item meaning. The finalized Urdu translation was approved for use in the main study.

### 2.4 Data analysis

Data was analyzed using SPSS v.29 (IBM, 2024) and R (Lavaan package). Initial analysis was carried out to assess the normality and descriptives of the MICA-4. Independent samples t-tests and one-way analysis of variance (ANOVA) were also used to examine differences in attitudes across demographic variables.

Item-level descriptive statistics (means, standard deviations, and skewness) were computed for both samples; corrected item-total correlations and “alpha if item deleted” were examined in Sample 1 to evaluate item discrimination and internal consistency.

Based on previously published factor structures of the MICA-4, we initially tested the original and alternative measurement models reported in prior validation studies. Because these models did not yield admissible solutions in our data, we proceeded with exploratory factor analysis to examine the underlying structure in this context.

We conducted EFA on Sample 1 to examine the underlying factor structure of the Urdu MICA-4 items. Principal axis factoring was used as the extraction method, with Promax rotation to allow correlated factors. Decisions regarding factor retention were based on eigenvalues, scree plot inspection, and interpretability of the factor solution.

We then performed CFA on Sample 2 to cross-validate the factor structure identified in Sample 1. Given the ordinal response format and non-normality, models were estimated using diagonally weighted least squares (DWLS). We evaluated the model fit using the Comparative Fit Index (CFI), Tucker-Lewis Index (TLI), Root Mean Square Error of Approximation (RMSEA), and Standardized Root Mean Square Residual (SRMR), using conventional cutoff criteria (e.g., CFI/TLI ≥ .90, SRMR ≤ .08–.10).

After establishing the final measurement model, we used Cronbach’s alpha and McDonald’s omega (composite reliability) to evaluated the internal consistency. We used average variance extracted (AVE) to assess convergent validity and the Heterotrait–Monotrait (HTMT) ratio to evaluate the discriminant validity.

## 3. RESULTS

This section presents our findings on demographics, reliability, and validation. Our demographic analyses revealed no significant difference across gender, education level, and experience levels (Table 1).

**Table 1:** Demographic characteristics of PCPs (sample 1 and sample 2)

| Table 1: Demographic characteristics of PCPs (sample 1 and sample 2) |  |  |  |  |
| --- | --- | --- | --- | --- |
| S. No. | Variable |  | N = 191 | N = 329 |
| 1 | Age | Mean (SD)= 31.91 (10.64) |  | Mean (SD) = 36.63 (8.25) |
| 2 | Gender | Male | 58.6 | .59.1 |
|  |  | Female | 41.4 | 40.9 |
| 3 | Education | Medical students | 30.4 | 76.8 |
|  |  | Medical graduates | 69.6 | 23.2 |
| 4 | Years of clinical experience | No experience | 29.8 | 0 |
|  |  | Less than 5 years | 17.3 | 40.2 |
|  |  | 5 to 10 Years | 15.7 | 34.1 |
|  |  | More than 10 years | 16.8 | 25.6 |
| 5 | Sector of work | Public | 56.5 | 81.7 |
|  |  | Private | 5.8 | 14.0 |
|  |  | Both | 17.3 | 0 |

### 3.1 Scale descriptives and reliability analysis

Table 2 presents the scale descriptives and reliability analysis of the scale. Item responses showed slight positive skewness, and the significant Shapiro–Wilk test indicated non-normality. The internal consistency estimate was modest (α = .64). Corrected item–total correlations suggested that removing Item 6R would increase alpha to .66; therefore, Item 6R was flagged for further evaluation in factor analysis.

**Table 2.** Descriptives of the scale.

| Table 2. Descriptives of the scale |  |  |  |  |  |  |  |
| --- | --- | --- | --- | --- | --- | --- | --- |
| Sample 1 (n = 191) |  |  |  |  |  | Sample 2 (n=329) |  |
| Items | Range Min-Max | M (SD) | Skewness | CITC | Cronbach's alpha if item deleted | M (SD) | Skewness |
| M1R | 1–6 | 3.66 (1.71) | -.142 | .240 | .625 | 3.17 (1.81) | 0.153 |
| M2R | 1–6 | 2.51 (1.60) | .768 | .370 | .605 | 2.39 (1.44) | 1.202 |
| M3 | 1–6 | 1.50 (1.22) | 2.852 | .162 | .634 | 1.39 (0.80) | 3.266 |
| M4R | 1–6 | 2.97 (1.71) | .351 | .376 | .603 | 2.92 (1.46) | 0.460 |
| M5R | 1–6 | 3.32 (1.54) | .136 | .417 | .599 | 4.19 (1.16) | -0.771 |
| M6R | 1–6 | 3.98 (1.60) | -.404 | .066 | .650 | 4.21 (1.44) | -0.589 |
| M7R | 1–6 | 3.43 (1.67) | -.034 | .229 | .627 | 3.23 (1.50) | 0.182 |
| M8R | 1–6 | 2.49 (1.59) | .733 | .331 | .611 | 2.43 (1.43) | 1.005 |
| M9 | 1–6 | 1.97 (1.67) | 1.634 | .130 | .642 | 2.39 (1.78) | 1.094 |
| M10 | 1–6 | 2.44 (1.52) | .923 | .287 | .618 | 2.47 (1.26) | 1.029 |
| M11 | 1–6 | 1.65 (1.14) | 2.163 | .242 | .625 | 1.72 (0.85) | 2.011 |
| M12 | 1–6 | 3.94 (1.61) | -.296 | .112 | .643 | 4.21 (1.31) | -0.678 |
| M13R | 1–6 | 2.58 (1.61) | .619 | .173 | .635 | 2.73 (1.45) | 0.594 |
| M14R | 1–6 | 3.58 (1.73) | -.132 | .219 | .628 | 3.31 (1.49) | 0.171 |
| M15R | 1–6 | 1.56 (1.21) | 2.244 | .346 | .613 | 1.93 (1.31) | 1.410 |
| M16 | 1–6 | 1.96 (1.33) | 1.596 | .373 | .608 | 2.12 (1.02) | 1.480 |

Note. R = reverse-scored items; SE= standard error; CITC = Corrected item total correlation.

### 3.2 Factor analysis

Initial confirmatory factor analyses testing the original five-factor model (Gabbidon et al., 2013), a previously reported three-factor model (Mısır et al., 2024; Vistorte et al., 2024), and a unidimensional model (Pereira, 2023), did not yield admissible solutions in our sample, likely due to model identification issues and unstable parameter estimates.

Therefore, we conducted EFA, where a five-factor solution resulted based on Kaiser criterion (eigenvalues > 1), different than the original study. The KMO measure of sampling adequacy was 0.673, indicating good adequacy of the sample for reliable factor extraction. Bartlett’s Test of Sphericity was significant (χ^2^ (120) = 575.78, p < .001), confirming that the correlations among variables were sufficient to proceed with factor analysis.

An initial five-factor solution emerged based on the Kaiser criterion (eigenvalues > 1); however, this solution lacked conceptual clarity and included cross-loadings. Item 6R demonstrated weak and inconsistent loadings across factors and was removed, supported by its low corrected item-total correlation and contribution to reduced internal consistency.

Following item removal, a three-factor solution was retained based on interpretability and simple structure. The final solution explained approximately 30% of the total variance. Factor loadings are presented in Table 3.

**Table 3:** Promax-rotated factor loadings from the Principal Axis Factoring of the MICA-4 Scale.

|  |  |  |  |  |  |
| --- | --- | --- | --- | --- | --- |
| Item no. | Items | Factor 1 | Factor 2 | Factor 3 | Uniqueness |
|  |  | Views | Stereotypes | Stigma |  |
| 3 | Working in the mental health field is just as respectable as other fields of health and social care. | 0.531 |  |  | 0.718 |
| 9 | If a senior colleague instructed me to treat people with a mental illness in a disrespectful manner, I would not follow their instructions. | 0.468 |  |  | 0.778 |
| 11 | I feel as comfortable talking to a person with a mental illness as I do talking to a person with a physical illness. | 0.758 |  |  | 0.430 |
| 15 | I would use the terms 'crazy', 'nutter', 'mad' etc. to describe to colleagues people with a mental illness who I have seen in my work. | **0.437** | 0.265 |  | 0.730 |
| 16 | If a colleague told me they had a mental illness, I would still want to work with them. | 0.615 |  |  | 0.585 |
| 10 | I feel as comfortable talking to a person with a mental illness as I do talking to a person with a physical illness. | **0.367** |  | 0.476 | 0.652 |
| 13 | If a person with a mental illness complained of physical symptoms (such as chest pain) I would attribute it to their mental illness. | 0.332 |  |  | 0.889 |
| 2 | People with a severe mental illness can never recover enough to have a good quality of life. |  | 0.464 |  | 0.740 |
| 4 | If I had a mental illness, I would never admit this to my friends because I would fear being treated differently. |  | 0.891 |  | 0.285 |
| 7 | If I had a mental illness, I would never admit this to my colleagues for fear of being treated differently. |  | 0.646 |  | 0.555 |
| 8 | Being a health/social care professional in the area of mental health is not like being a real health/social care professional. |  | 0.336 |  | 0.835 |
| 14 | General practitioners should not be expected to complete a thorough assessment for people with psychiatric symptoms because they can be referred to a psychiatrist. |  | 0.292 |  | 0.827 |
| 1 | I just learn about mental health when I have to, and would not bother reading additional material on it. |  |  | 0.427 | 0.745 |
| 5 | People with a severe mental illness are dangerous more often than not. |  |  | 0.431 | 0.687 |
| 12 | The public does not need to be protected from people with a severe mental illness. |  |  | 0.494 | 0.786 |

Factor 1 (Views) included seven items (3, 9, 11, 15, 16, 10, 13) reflecting clinicians’ general evaluative perspectives toward mental illness and professional roles. It has items related to considering mental health as a respectable profession, somatic attribution, physicians checking psychological health with physical health, being comfortable talking to, or working with people with ill mental health. This subscale refers to concepts that can be based on personal beliefs, perceptions, experiences, or knowledge.

Factor 2 (stereotypes) included five items (2, 4, 7, 8, 14) representing generalized beliefs and disclosure-related concerns regarding individuals with mental illness. This contains items such as people with mental illness can never recover, have a good quality of life, fear of disclosure to friends and colleagues, and mental health professionals are not real professionals. Stereotypes refer to generalized and fixed beliefs about a group, which can be positive or negative.

Factor 3 (Stigma) included three items (1, 5, 12) capturing social distancing and perceived threat-related attitudes. Stigma, which is a form of social disapproval or disgrace attached to a particular characteristic, often leading to discrimination. Stigma is the negative reaction and consequences that follow stereotypes. This includes items such as people with mental illness are dangerous, the public should be protected from them, and reading about mental illness only when required.

### 3.3 Confirmatory factor analysis

The three-factor model demonstrated acceptable overall fit (Table 4). The Comparative Fit Index (CFI = .958) indicated good fit, and the Tucker–Lewis Index (TLI = .946) indicated acceptable fit. The Standardized Root Mean Square Residual (SRMR = .085) fell within recommended thresholds for ordinal models. The Root Mean Square Error of Approximation (RMSEA = .108) was elevated and was interpreted with caution. Prior research suggests that RMSEA may be inflated when using the Diagonally Weighted Least Squares (DWLS) estimator with ordinal indicators and limited degrees of freedom (Kenny et al., 2015). Two alternative two-factor models were tested to evaluate structural parsimony. A model combining Views and Stigma demonstrated slightly poorer fit (CFI = .946, TLI = .932, SRMR = .088) than the three-factor model. A second model combining Views and Stereotypes showed further reduction in fit (CFI = .942, TLI = .927, RMSEA = .112). Collectively, these comparisons support retention of the three-factor structure.

**Table 4:** Summary of model fit indices.

| Summary of the model fit Indices |  |  |  |  |  |  |
| --- | --- | --- | --- | --- | --- | --- |
| | $\chi^2$ | Df | CFI | TLI | SRMR | RMSEA |
| **3-Factor Model** | 244.73 | 51 | 0.958 | 0.946 | 0.085 | 0.108 |

Based on standardized factor loadings, three items (Q9, Q13, and Q12) were removed due to weak loadings (< .40). The final model was estimated using the DWLS estimator, appropriate for ordinal Likert-type data. Composite reliability (ω) indicated adequate internal consistency for the Views (ω = .70) and Stereotypes (ω = .74) factors. The Stigma factor demonstrated lower reliability (ω ≈ .53), likely reflecting its two-item structure. Convergent validity was modest, with average variance extracted (AVE) values ranging from approximately .40 to .44.

The factor loadings, composite reliability, and average variance extracted, assessing the strength and reliability of each construct, are presented in *supplementary material B*.

Although the correlation between the Views and Stereotypes factors was high (r = .86), all HTMT values were below recommended thresholds, indicating that the constructs remain empirically distinct (see *supplementary material C*).

## DISCUSSION

This study tested reliability and validity of an Urdu translation of MICA-4, and clarified its factor structure for use in Pakistan. We avoided substantive modification of item content, as advised by the original developers, and carefully translated the scale to retain meaning and intent as closely as possible. Minor linguistic refinements were made to improve clarity and comprehension in the local context while preserving conceptual equivalence. These refinements emphasized widely understood terminology and, where appropriate, inclusion of brief examples to support interpretation without changing the intended meaning of the items. For instance, item 10 was translated with brief parenthetical examples (e.g., depression; diabetes) for effective interpretation by the PCPs without changing the underlying construct. Likewise, item 8 was rephrased to reflect language commonly used in clinical settings while retaining the item’s intended meaning.

We did not find any significant differences in attitudes across PCPs’ demographics, including gender, age, and years of experience, although Gabbidon et al., (2013) and Ghuloum et al., (2022) had reported that female doctors and Vistorte et al., (2018) found that younger and less experienced doctors tend to exhibit less stigmatizing attitudes compared to older and more experienced clinicians. Our finding can be attributed to pervasive stigma, gaps in psychiatric training during medical education, and inadequate mental health literacy among healthcare professionals in Pakistan (Husain et al., 2020; Khan et al., 2023).

The original five-factor structure of the scale demonstrated by Gabbidon et al., (2013), has not been supported by other validation studies. The validation study in the Spanish and Portuguese (Vistorte et al., 2023), and Turkish population (Mısır et al., 2024) has demonstrated a three-factor structure. Whilst a study in Brazil reported a two-factor structure (Fernandes et al., 2024), and another study on Portuguese population demonstrated a unifactorial nature of the scale (Sofia & Pereira, 2023). This variability across studies suggests that the dimensionality of the MICA-4 may be sensitive to contextual and cultural factors rather than reflecting a universally stable structure. We also agree with Bizumic et al., (2022) that measures to assess stigma towards mental illness have certain methodological issues, like lacking psychometric rigor, poor validity, non-replicable factor structures, and complex language.

Our study reinforces the complexity of validating the MICA-4 across cultural contexts. Internal consistency in our first sample was modest, a pattern already reported in other validation studies (Fernandes et al., 2024; Sofia & Pereira, 2023). Certain items demonstrated weak loadings and limited discrimination, particularly Item 6, which Mısır et al (2024) had also removed due to consistently low factor loadings and conceptual ambiguity in this context. We also found that items 9, 12, and 13 did not demonstrate adequate performance in the confirmatory model and had to exclude these to achieve a more stable structure. The resulting model demonstrated improved reliability and structural clarity. Other validation studies have reported item-level inconsistencies and structural modifications to improve model fit (Fernandes et al., 2024; Mısır et al., 2024; Sofia & Pereira, 2023). We agree that limited flexibility in item refinement or removal may be necessary to achieve psychometric stability across diverse settings. Allowing structured, evidence-based refinement while preserving conceptual equivalence may enhance the scale’s cross-cultural applicability without compromising its theoretical foundation. The variability reported across validation studies also raises a practical implication for scale dissemination. When item-level instability repeatedly emerges across contexts, permitting limited, evidence-based refinement may strengthen measurement stability and comparability, as well as its reliability in diverse healthcare settings.

Our findings supported a three-factor structure, namely views, stereotypes and stigma. In our case, both items 4 and 7 have strong factor loadings on stereotype, indicating that the fear of telling friends or family about mental illness is due to a strongly negative societal belief. Lack of support from family members and opposition to seek medical help also contributes to a preference for non-disclosure (Samari et al., 2022). A strong reluctance to seek treatment for mental health issues due to fear of stigma and negative career repercussions, even among HCPs, is also known (Braquehais & Vargas-Cáceres, 2023; Mehta & Edwards, 2018). Items 4 and 7 always pair in the same factor in all cultural settings. However, in English and Latin American version, it is in the factor structure “disclosure” versions (Gabbidon et al., 2013; Vistorte et al., 2023); whereas in the Turkish version, it is attributed to social distance (Mısır et al., 2024).

We agree with Corrigan and colleagues (2014) that stigma is a complex construct that includes public, self, and structural components. It directly affects people with mental illness, as well as their support system, provider network, and community resources. The effects of stigma are moderated by knowledge of mental illness and cultural relevance. Understanding stigma is central to reducing its negative impact on care seeking and treatment engagement (Corrigan et al., 2014).

Unlike Mısır et al., (2024) who reported that people with mental illness are perceived as dangerous because of stereotyping, we conclude that it is likely to be a result of pervasive public and systemic stigma. Even in our context, mental illness is often perceived as a sign of weak faith, demonic possessions, divine punishment, or a result of moral transgression - causing social distancing and perceiving them as dangerous; the stigma can be moderated by mental health literacy and awareness (Ahad et al., 2023; Corrigan et al., 2014; Corrigan & Watson, 2002; Mansoor & Warsi, 2023).

The psychometric evaluation of the scale via CFA highlights the underlying structure of the constructs. The high latent correlation between views and stereotypes suggests a significant conceptual overlap, indicating that participants’ cognitive beliefs (stereotypes) are closely tied to their broader evaluative perspectives (views). However, the superior fit of the three-factor model over the collapsed two-factor model, coupled with the acceptable HTMT ratio, confirms that these factors remain empirically distinct. This distinction is important for future interventions: while stereotypes and views are closely related, they may require different strategies, one targeting factual misconceptions and the other addressing broader negative attitudes. The lower reliability of the stigma factor is likely due to the presence of only two-items. While this factor offers initial insights into social distancing, future versions of the instrument should consider expanding this subscale to improve internal consistency and capture a broader range of stigmatizing behaviors.

We observed a pattern, well noted previously, that items 3, 9, 10, 11, and 16 consistently load on a similar underlying dimension across multiple validation studies (Gabbidon et al., 2013; Mısır et al., 2024; Vistorte et al., 2023). However, these items were named as views of health/social care field and mental illness or patient-care for people with mental illness in other studies. Differences in factor labeling are likely to be a reflection of contextual interpretations rather than substantive differences in item meaning, suggesting that clinicians’ perceptions of mental illness are shaped by sociocultural influence and healthcare system. For instance, Fernandez et al., (2024) defined the dimensionality of the scale as views that are divided into two factors, as either views on mental healthcare or on people with mental illness.

As reported by Misir and colleagues (2024), Item 15 represents devaluation during patient-care. Such derogatory terminology is recognizable within local clinical discourse, reflecting entrenched negative beliefs about individuals with mental illness. In our study, this item loaded on views and stereotypes, indicating that stigmatizing language may simultaneously reflect attitudinal beliefs and professional positioning. The literature also supports that not only the community members but also PCPs have negative and pejorative attitudes towards people with mental illness, which deter patients from seeking professional help (Haddad et al., 2016).

One Item regarding protection from people with severe mental disorders (Item 12) is in the factor “views of healthcare field and mental illness” in English and Latin American versions (Gabbidon et al., 2013; Vistorte et al., 2023), while named as “social distance of healthcare professionals from people with mental illness” in the Turkish version (Mısır et al., 2024). In our study, its loading pattern suggested closer conceptual alignment with stigma-related beliefs, as stigma often manifests through fear-based reactions, discrimination, or social disapproval (Corrigan et al., 2014).

Like other reports, our CFA findings support the context-sensitive nature of the MICA-4 structure. The final 12-item version, excluding items 6, 9, 12, and 13, is found to be more reliable and deemed relevant to our cultural and healthcare context. We recommend that the Urdu MICA-4 be used with consideration of its measurement characteristics within this context. The three-factor structure demonstrated acceptable model fit and construct validity; however, internal consistency estimates were modest in Sample 1. This pattern is consistent with existing literature, which reports variability in reliability across cultural settings (Fernandes et al., 2024).

The sample size in our study may limit generalizability, but non-probability sampling is widely accepted in early-stage validation research, particularly when the primary aim is psychometric evaluation (Kalton, 2023). Our study did not include an assessment of convergent or divergent validity, which would be valuable in determining whether MICA-4 captures stigmatising attitudes consistent with other established tools.

## Conclusion

This study highlights the importance of local validation and suggests that limited, evidence-based refinement can enhance the application of MICA-4 without compromising conceptual equivalence. Further validation in diverse healthcare samples may strengthen the generalizability of the Urdu MICA-4 across clinical settings in Pakistan.

## Authors Contribution

NM: Conceptualization, Data collection, data curation, writing – original draft

AsN: Methodology, formal analysis, Writing – original draft

AH: Visualization, Supervision, Project administration, Writing – review and editing.

## Acknowledgments

We would like to thank Dr. Arooj Najmussaqib for her contribution to methodology in an initial draft.

## Ethical Statement

This study was conducted as part of the Mental Health and Psychosocial Support Project. The ethical approval for this study was granted by the review committee of the Health Section at the Ministry of Planning, Development & Special Initiatives, in compliance with ethical standards and consent protocols under letter no. 6(262) HPC/2020.

## Availability of data and materials

The aggregate data that support the findings of this study can be made available upon request from the Mental Health Strategic Planning & Coordination Unit, at the Health Section of the Ministry of Planning, Development & Special Initiatives, Government of Pakistan.

## Patient and Public Involvement

Patients or the public were not involved in the design, conduct, reporting, or dissemination plans of our research.

## Consent for publication

Not applicable.

## List of abbreviations

LMIC: Low- and middle-income countries
HCPs: Healthcare Providers
PCPs: Primary care physicians
MICA-4: Mental Illness Clinicians’ Attitudes Scale, version 4
CAMI: Community Attitudes Toward the Mentally Ill
OMI: Opinions About Mental Illness

## Supplementary material A: Inclusion and exclusion criteria of PCPs

Inclusion criteria were: (1) currently practicing PCPs in Pakistan, (2) holding a medical degree (MBBS or equivalent). These criteria required participants to hold a recognized professional qualification, demonstrate interest in professional development, show an inclination toward a biopsychosocial approach to care, and adhere to ethical standards.

Exclusion criteria included enrollment in any specialization (psychiatry or mental health programme), or mental health professionals (e.g., psychiatrists, psychologists).

## Supplementary material B: Measurement model properties

| Factor and Items | Standardized Loading | p-value | $\omega$ (CR) | AVE |
| --- | --- | --- | --- | --- |
| **Views** |  | < .001 | **0.70** | **0.41** |
| Q3 | 0.622 | --- |  |  |
| Q11 | 0.542 | < .001 |  |  |
| Q15 | 0.883 | < .001 |  |  |
| Q16 | 0.599 | < .001 |  |  |
| Q10 | 0.478 | < .001 |  |  |
| **Stereotypes** |  | < .001 | **0.76** | **0.44** |
| Q2 | 0.665 | --- |  |  |
| Q4 | 0.707 | < .001 |  |  |
| Q7 | 0.653 | < .001 |  |  |
| Q8 | 0.643 | < .001 |  |  |
| Q14 | 0.658 | < .001 |  |  |
| **Stigma** |  | < .001 | **0.55** | **0.40** |
| Q1 | 0.755 | --- |  |  |
| Q5 | 0.481 | < .001 |  |  |

**Note:** $\omega$ = McDonald's Omega (CR = Composite Reliability); AVE = Average Variance Extracted. For the first item of each factor, the loading is fixed to 1.0 in the unstandardized model, so no p-value is calculated.

## Supplementary material C: Discriminant validity using the HTMT ratio

**Table S2:** Discriminant validity using the HTMT ratio.

| Table S2: Discriminant validity using the HTMT ratio |  |  |  |
| --- | --- | --- | --- |
| Factor | 1. Views | 2. Stereotypes | 3. Stigma |
| **1. Views** | - |  |  |
| **2. Stereotypes** | **0.827** | - |  |
| **3. Stigma** | **0.672** | **0.786** | - |

**Note:** Values less than 0.90 indicate acceptable discriminant validity.

## Footnotes

1 https://www.indigo-group.org/wp-content/uploads/2024/01/Translations.pdf

